# COVID-19: a crash test for biomedical publishing?

**DOI:** 10.1101/2020.06.13.20130310

**Authors:** Ivan Y Iourov, Maria A Zelenova, Svetlana G Vorsanova

## Abstract

The effect of COVID-19 on biomedical publishing (BP) (i.e. scientific biomedical periodicals continuously published by research communities or commercial publishers) has not been deeply explored. To estimate the immediate COVID-19 impact on BP, we have assessed <u>P</u>ub<u>M</u>ed-<u>i</u>ndexed <u>a</u>rticles about <u>C</u>OVID-19 (PMIAC) from December 2019 to April 2020. PMIAC have been classified according to publication date, country, and journals for evaluation of time-, region- and scientometric-dependant impact of COVID-19 on BP and have been curated manually (i.e. each entry has been individually analyzed). PMIAC analysis reflects geographic and temporal parameters of outbreak spread. A major BP problem is related to the fact that only 40% of articles report/review/analyze data. Another BP weakness is the clusterization of “highly-trusted” publications according to countries of origin and “highly impacting” journals. Finally, a problem highlighted by COVID-19 crisis is the increased specification of biomedical research. To solve the problem, analytical reviews integrating data from different areas of biology and medicine are required. The data on PMIAC suggest priority of “what is published” over “where it is published” and “who are the authors”. We believe that our brief analysis may help to shape forthcoming BP to become more effective in solving immediate problems resulted from global threats.

---

The novel coronavirus disease (COVID-19) has become an unprecedented challenge for biomedical research and practice.^1,2^ COVID-19 is currently designated as the cause of pandemics and world-wide crisis, which has profound effects on all areas of life.^3^ Due to the global nature, the extent of COVID-19 impact has not been evaluated in a variety of important fields. For instance, the effect of COVID-19 on biomedical publishing (BP) (i.e. scientific biomedical periodicals continuously published by research communities or commercial publishers) has not been deeply explored. COVID-19 crisis has to affect BP as the unique source of evidence-based knowledge in healthcare, medicine and the life sciences. Accordingly, BP perspectives in the post-COVID-19 era seem to be important for biomedical researchers and practitioners around the world. To estimate the immediate COVID-19 impact on BP, we have assessed <u>P</u>ub<u>M</u>ed-<u>i</u>ndexed <u>a</u>rticles about <u>C</u>OVID-19 (PMIAC) from December 2019 to April 2020.

Articles retrieved from PubMed (from 1 December 2019 to 30 April 2020) have been classified according to publication date, country, and journals for evaluation of time-, region- and scientometric-dependant impact of COVID-19 on BP. Retrieved articles have been curated manually (i.e. each entry has been individually analyzed) to limit the number of articles to those fitting the format of original research, review, case-report, meta-analysis, hypothesis, or systematic review. News, comments, “comments to comments” and similar article types have been excluded. Additionally, articles’ number has been limited by exclusion of PMIAC by international collaborations for a fairer analysis of country performances. To address scientometric parameters of COVID-19 publications, we have correlated journal’s impact factors (IF) with number of articles (n≥2). Regional distribution and performance in BP dedicated to COVID-19 have been compared to Nature Index (excellence in “high quality” BP)^4^ and Scimago Journal & Country Rank (excellence in global peer-reviewed BP).^5^

Generally, time-dependent increase of PMIAC numbers has correlated with the crisis progression. In 2019, there were 4 PMIAC in contrast to 6793 and 5899 PMIAC (retrieved tags: [dp] (Date of Publication) and [CRDT] (Create Date), respectively) during the first 4 months of 2020. After manual curation, the amount of [CRDT] PMIAC has been narrowed to 2362 (Fig. 1A). Exclusion of international collaborative PMIAC limited the amount to 1402. As expected, number of PMIAC increased from month to month as the crisis deepened (Fig. 1B). Addressing scientometric segregation of PMIAC among journals in January and February 2020 has been made using the most recent IF (taken from journal’s web sites). PMIAC have been clearly distributed among following IF categories: January — i) high-impact (IF>40); ii) low-to-average-impact (IF=0-10) (Fig. 1C); February — i) high-impact (IF>40); ii) high-to-average-impact (IF=10-30); iii) low-to-average-impact (IF=0-10) (Fig. 1D). Each impact-factor-dependent cluster has demonstrated comparable PMIAC numbers. Country distribution and excellence performance according to analysis of PMIAC has demonstrated that “key players” (top ~10 countries) are almost the same as those defined by Nature Index and Scimago Journal & Country Rank. However, the leadership in PMIAC is different as to scientometric rankings (Fig. 1E).

**Figure 1.**
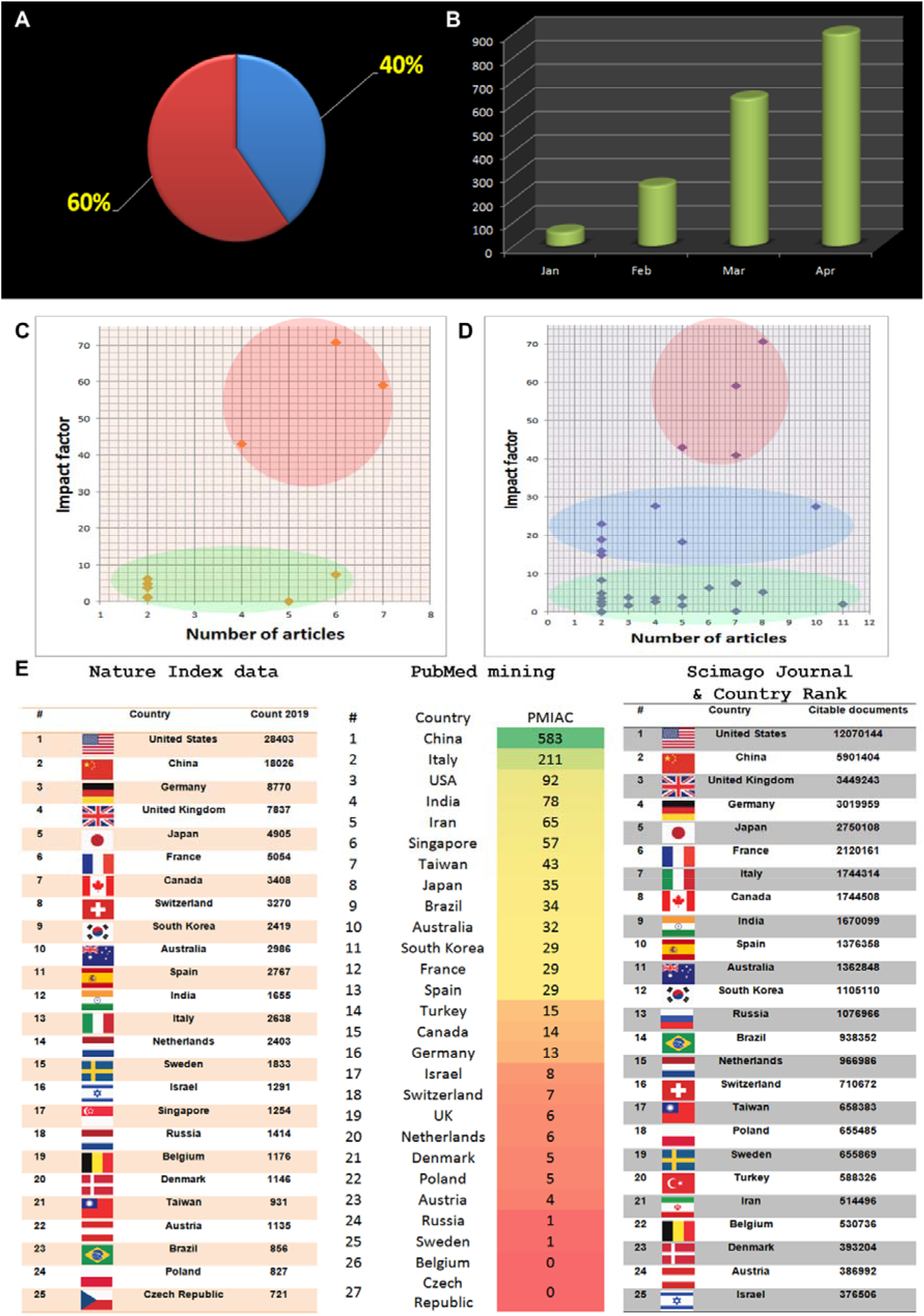
(A) Percentage of PMIAC curated manually: PMIAC reporting and reviewing data (n=2362) — 40%, the remainder — 60%. (B) Increase of PMIAC numbers month by month. (C and D) PMIAC numbers in function of journals’ IF in January (C) and February (D). (E) Country’s leadership in BP according to Nature Index, our COVID-19 PMIAC analysis (PubMed search: COVID-19 + Country) excluding articles by extended international collaborations and Scimago journal & country rank.

Certainly, PMIAC analysis reflects geographic and temporal parameters of outbreak spread. Nonetheless, bibliographic analysis highlights several important aspects of current BP, which might be important for its shaping in the post-COVID-19 era. Figuratively, COVID-19 effect on BP is like a crash test, which explicitly demonstrates weaknesses of a vehicle by one disastrous event. The explosive rise of PMIAC shows the shock effect caused by a drastic outbreak. A major BP problem is related to the fact that only 40% of articles report/review/analyze data (Fig. 1A), whereas the remainder does not really contribute to biomedical research *per se*. Another BP weakness is the clusterization of “highly-trusted” publications according to countries of origin and “highly impacting” journals. Since a crisis may start in any part of the world, such kind of clusterization causes inability of global research community to respond immediately/adequately to the challenges. To publish the data as quick as possible for helping researchers and physicians world-wide, authors prefer to submit their articles ignoring scientometrics. Publication prestige is sacrificed for the emergency. Disastrous spread of COVID-19 has resulted in acknowledgement that all data matter and has urged researchers/physicians combating the crisis to ignore publishing stereotypes. In these circumstances, the level of expertise (i.e. presence/lack of peer-reviewing) is not a priority. The most important data and experience were initially provided by researchers on the cutting edge, i.e. Chinese colleagues.^6,7^ At the international level, understating of emergent action in biomedical research of COVID-19 has led to a recognition of an need for concerned action of internationally integrated researchers.^8^ Unfortunately, this recognition has not resulted in large-scale analyzing of BP efficiency neither during pandemics nor for the post-COVID-19 era. Nevertheless, constructive criticism of current BP paradigm applicable to sharing knowledge about specific aspects of COVID-19 is available in biomedical literature.^9^ Fortunately, the paradigm of open science has been widely introduced and has encompassed almost all COVID-19 publications.^10^ Still, there remains a concern about scientific level and merit of publications appeared immediately due to pandemics’ crisis. On the other hand, it does not necessarily mean that “highly-trusted” publications in “highly-impacting” journals guarantee the intrinsic applicability and usefulness for further research and practice. COVID-19 test of BP clearly indicates that there is a need for immediate assessment of publishable research results, which current scientometrics is unable to do. This suggests priority of “what is published” over “where it is published” and “who are the authors”. Although this idea is consistently highlighted, general researchers’ experience does not confirm that it is absolutely true. COVID-19 crisis is an promising opportunity to make BP more applicable for solving actual problems in biomedicine, taking into account the state-of-art in information management and distribution. For instance, there have been extended discussions about the requirements of new models of BP during the last years. These models are generally based on open-access, preprinting, social-media-like feedback, and preregistration. COVID-19 crisis gives further empirical support to such discussions shaping BP future. It is highly likely that peer review and manuscript/data curation may precede the ultimate publication of article in an open-access manner.^11,12^ Here, one can be aware about a limitation produced by current open-access system, i.e. the inability of the majority of researchers around the world to pay article processing charges. Certainly, this problem is generally solved for each individual manuscript, but it takes appreciable time, which is so precious during outbreak spread. Finally, a problem highlighted by COVID-19 crisis is the increased specification of biomedical research.^3,9^ Accordingly, each PMIAC represents a single “gem” reporting data on extremely specific aspects of COVID-19, but analytical “corona-reviews”, which systematically analyze these data to produce an integrated view for COVID-19 treatment and prevention, are rare. This is not only actual for PMIAC, but for BP, as a whole. Therefore, analytical reviews integrating data from different areas of biology and medicine are required. In total, it appears that BP should evolve more rapidly and efficiently after COVID-19 crisis test. This evolution would be productive based on actual epistemological concepts (e.g. analytical transdisciplinary reviews) and opportunities offered by modern information technologies.

The consequences of COVID-19 pandemics are only starting to be evaluated. Without deep analysis, unsolved problems produced by COVID-19 pandemics may have long-term deleterious effects. Challenges created by COVID-19 require changes in various dimensions of biomedical research including BP. We believe that our brief analysis may help to shape forthcoming BP to become more effective in solving immediate problems resulted from global threats.

## Data Availability

All the data is provided in the manuscript.

## ACKNOWLEDGMENTS

Authors are partially supported by RFBR and CITMA according to the research project No. 18-515-34005. Prof. IY Iourov is supported by the Government Assignment of the Russian Ministry of Science and Higher Education, Assignment no. AAAA-A19-119040490101-6. Prof. SG Vorsanova is supported by the Government Assignment of the Russian Ministry of Health, Assignment no. AAAA-A18-118051590122-7.

## AUTHOR CONTRIBUTIONS

IYI analyzed data and wrote the manuscript. MAZ analyzed data. SGV made significant theoretic contribution.

### Competing interests

The authors declare that they have no competing interests.

## Notes

### Competing Interest Statement

The authors have declared no competing interest.

### Clinical Trial

Not applicable

### Author Declarations

This is the analysis of bibliographic data published in peer-reviewed periodicals indexed in publicly accessible databases. No patients are involved in this study. Accordingly, there is no need for specific clinical trial registration, ethics committee approvals or related actions.

